# Hair loss in females and thromboembolism in males are significantly enriched in post-acute sequelae of COVID (PASC) relative to recent medical history

**DOI:** 10.1101/2021.01.03.20248997

**Authors:** Elliot Akama-Garren, Praveen Anand, Tudor Cristea-Platon, Patrick Lenehan, Emily Lindemer, Sairam Bade, Saran Liukasemsarn, John C. O’Horo, Ryan T. Hurt, Amy W. Williams, Gregory J. Gores, Andrew D. Badley, Samir Awasthi, Venky Soundararajan

**Affiliations:** nference, inc., One Main Street, Suite 400, East Arcade, Cambridge, MA 02142, USA; nference Labs,2nd Floor, 22 3rd Cross Rd, Murgesh Pallya, Bengaluru, India; Mayo Clinic, Rochester MN, USA

## Abstract

After one year of the COVID-19 pandemic, over 130 million individuals worldwide have been infected with the novel coronavirus, yet the post-acute sequelae of COVID-19 (PASC), also referred to as the ‘long COVID’ syndrome, remains mostly uncharacterized. We leveraged machine-augmented curation of the physician notes from electronic health records (EHRs) across the multi-state Mayo Clinic health system to retrospectively contrast the occurrence of symptoms and diseases in COVID-19 patients in the post-COVID period relative to the pre-COVID period (n=6,413). Through comparison of the frequency of 10,039 signs and symptoms before and after diagnosis, we identified an increase in hypertensive chronic kidney disease (OR 47.3, 95% CI 23.9-93.6, p=3.50×10^−9^), thromboembolism (OR 3.84, 95% CI 3.22-4.57, p=1.18×10^−4^), and hair loss (OR 2.44, 95% CI 2.15-2.76, p=8.46×10^−3^) in COVID-19 patients three to six months after diagnosis. The sequelae associated with long COVID were notably different among male vs female patients and patients above vs under 55 years old, with the hair loss enrichment found primarily in females and the thromboembolism enrichment in males. These findings compel targeted investigations into what may be persistent dermatologic, cardiovascular, and coagulopathic phenotypes following SARS-CoV-2 infection.

## Introduction

Since its emergence in December 2019, severe acute respiratory syndrome coronavirus 2 (SARS-CoV-2) and its clinical syndrome coronavirus disease 2019 (COVID-19) have elicited clinical presentations ranging from asymptomatic disease to severe respiratory failure and death, as well as a number of extra-pulmonary manifestations. Despite extensive characterization of its acute presentation, relatively little is known about the long-term effects of both mild and severe COVID-19^1,2^. Telephone interviews of outpatient SARS-CoV-2 infected patients 14-21 days after positive RT-PCR test identified persistence of fatigue, cough, dyspnea, and headache most commonly^3^. A study of 143 recovered COVID-19 patients 60 days after symptom onset found that 87.4% still had at least one symptom, most commonly fatigue or dyspnea^4^. Another study of 55 recovered COVID-19 patients 3 months after discharge found 71% had radiologic abnormalities and 25% had abnormal pulmonary function tests^5^.

A better understanding of the convalescent phase and long-term consequences of COVID-19 (“long COVID”) is crucial as the number of patients who have been infected with SARS-CoV-2 and recover from COVID-19 continues to rise. Several efforts are ongoing to monitor self-reported symptoms following COVID-19 recovery^6^, but results from these studies will be limited by the pace at which patients are recruited. Current attempts to assess the long-term sequelae of COVID-19 using structured data such as International Classification of Disease (ICD) codes are limited by a finite set of possible variables and are biased by *a priori* selection of variables of interest.

We have previously reported using an augmented curation approach, wherein BERT-based neural networks are used to identify diseases and symptoms that have been positively attributed to patients in unstructured clinical documentation to curate millions of notes in a short period of time^7^. Here we leverage augmented curation to retrospectively curate the complete electronic health records (EHRs) of COVID-19 patients in order to identify symptoms and phenotypes that persist several months after diagnosis of the viral infection.

## Results

To identify signs and symptoms associated with long COVID, we performed augmented curation from clinical notes of COVID-19 patients 1-6 months before and 3-6 months after diagnosis. Using deep neural networks over an institution-wide platform, we extracted positively attributed signs, symptoms, and diseases from Mayo Clinic electronic health records (EHRs). Among 10,039 signs and symptoms identified from the EHRs of 6,413 COVID-19 patients (Table 1), we identified seven terms significantly increased after diagnosis relative to before diagnosis: infection (OR 1.72, 95% CI 1.67-1.78, p=1.31×10^−29^), pneumonia (OR 1.91, 95% CI 1.81-2.01, p=1.15×10^−14^), hypertensive chronic kidney disease (OR 47.3, 95% CI 23.9-93.6, p=3.50×10^−9^), thromboembolism (OR 3.84, 95% CI 3.22-4.57, p=1.18×10^−4^), venous thromboembolism (OR 5.91, 95% CI 3.22-3.84, p=1.84×10^−4^), and hair loss (OR 2.44, 95% CI 2.15-2.76, p=8.45×10^−3^), and immunodeficiency (OR 4.13, 95% CI 3.31-5.16, p=1.95×10^−2^) (Figure 1, Supplementary Table 1). In contrast, 183 terms were more prevalent before diagnosis, most significantly pain (OR 0.65, 95% CI 0.63-0.66, p=5.14×10^−39^), hypertension (OR 0.67, 95% CI 0.65-0.68, p=1.26×10^−36^), and swelling (OR 0.58, 95% CI 0.56-0.60, p=8.75×10^−28^). These findings suggest that hypertensive chronic kidney disease, coagulopathies, and hair loss might be associated with long COVID.

**Table 1:**
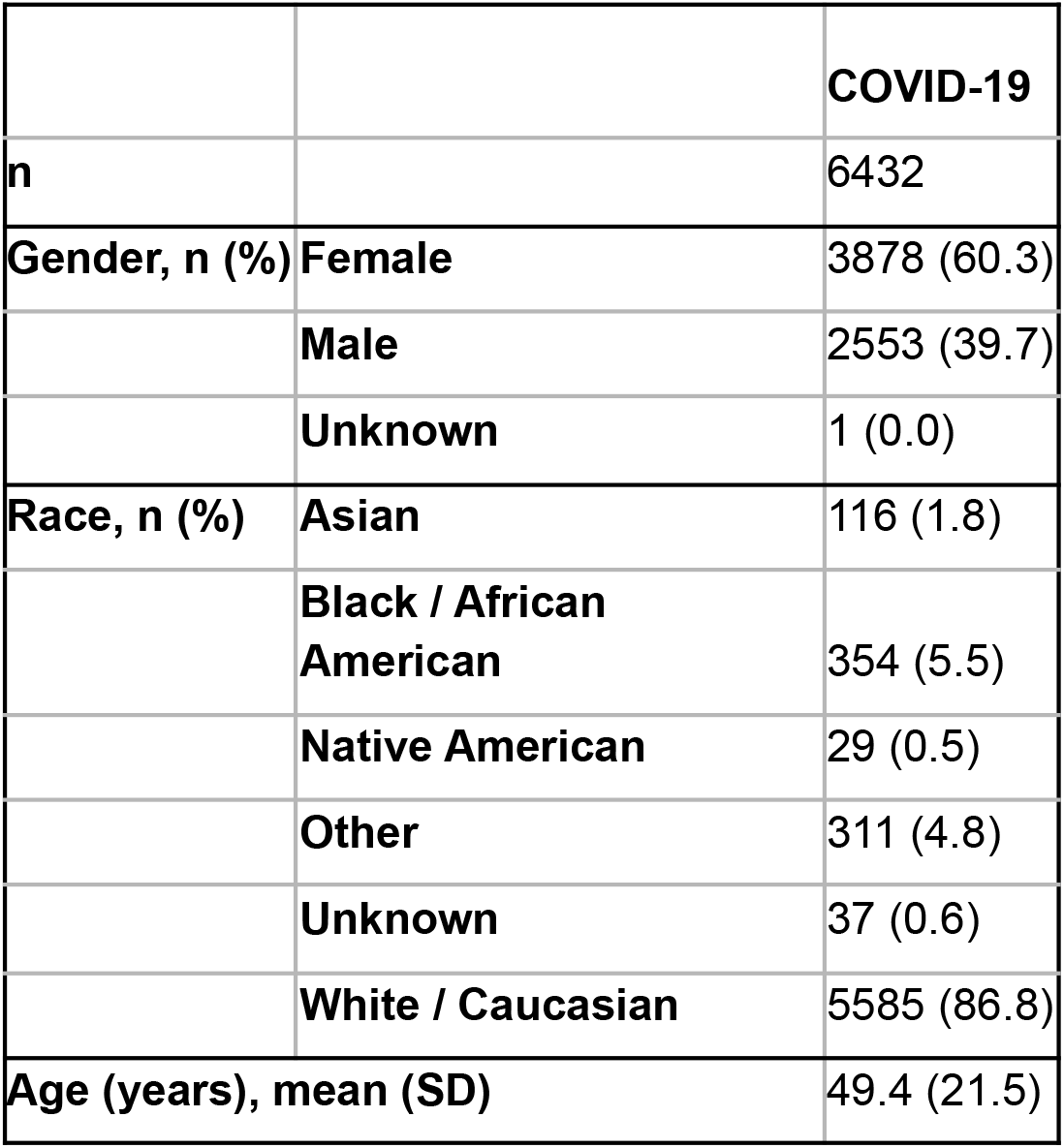
Demographics of the COVID-19 cohort analyzed in this study.

|  |  | **COVID-19** |
| --- | --- | --- |
| **n** |  | 6432 |
| **Gender, n (%)** | **Female** | 3878 (60.3) |
|  | **Male** | 2553 (39.7) |
|  | **Unknown** | 1 (0.0) |
| **Race, n (%)** | **Asian** | 116 (1.8) |
|  | **Black / African American** | 354 (5.5) |
|  | **Native American** | 29 (0.5) |
|  | **Other** | 311 (4.8) |
|  | **Unknown** | 37 (0.6) |
|  | **White / Caucasian** | 5585 (86.8) |
| **Age (years), mean (SD)** |  | 49.4 (21.5) |

**Figure 1.**
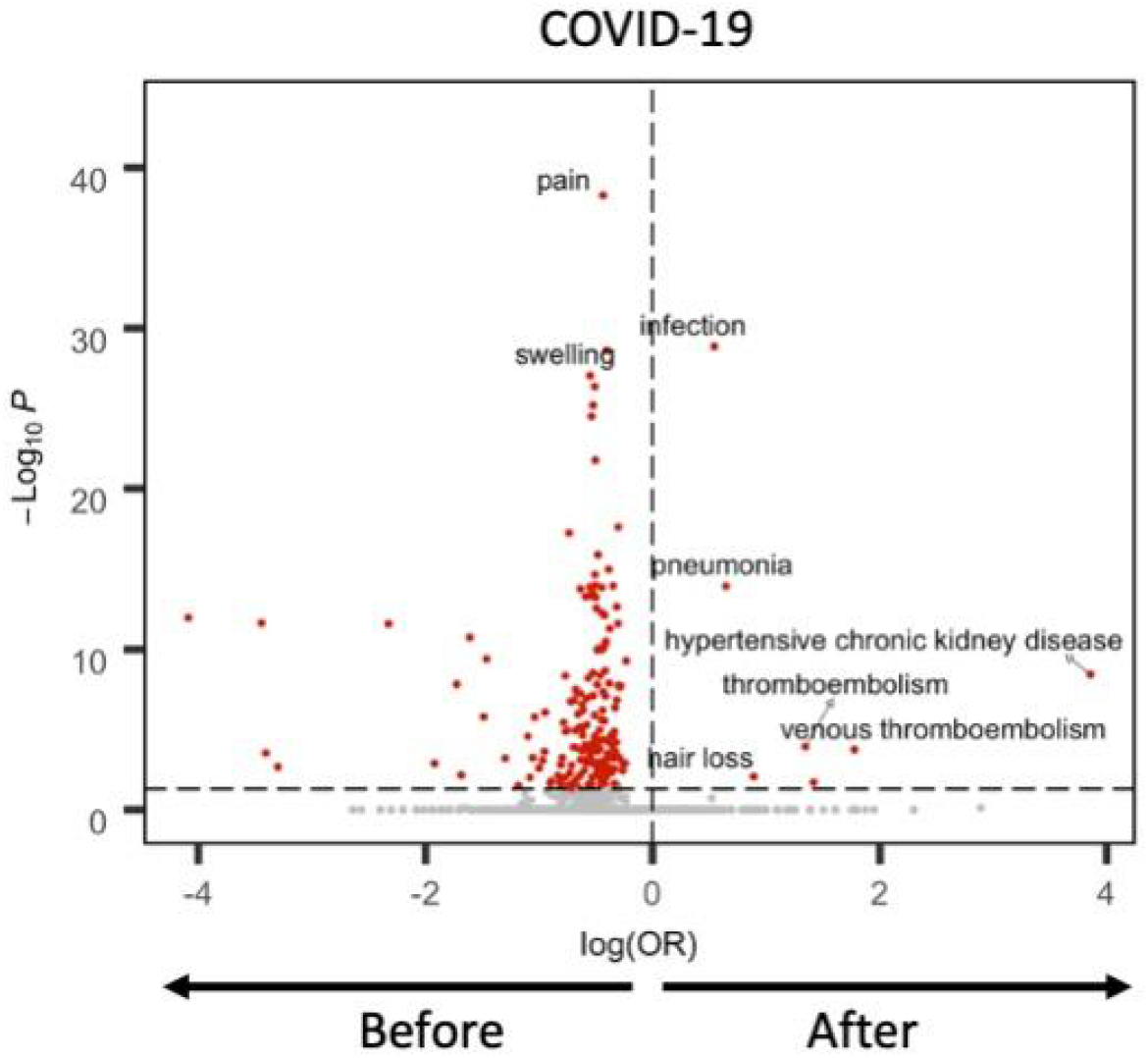
Coagulopathies are associated with long COVID. Volcano plot displaying odds ratios (OR) of the change in prevalence before vs after disease for each sign and symptom identified from patient charts of COVID-19 patients. P-values calculated by McNemar test and adjusted for multiple comparisons using Bonferroni correction. Significant (p<0.05) signs and symptoms indicated in red.

Demographic and clinical stratification of patients revealed distinct signs and symptoms associated with long COVID (Table 2). For female patients (n=3,870), only infection and hair loss were significantly enriched after COVID-19 diagnosis, while infection, pneumonia, and thromboembolism were specifically enriched among male patients in the post-COVID period (Figure 2). For patients under 55 years old (n=3,464), only infection was significantly enriched after COVID-19 diagnosis (Figure 3). Among hospitalized (n=703) and ICU-admitted (n=209) patients, pneumonia and respiratory failure were documented more frequently in clinical notes 3-6 months after COVID-19 diagnosis than in the 1-6 months before (Figure 4).

**Table 2:**
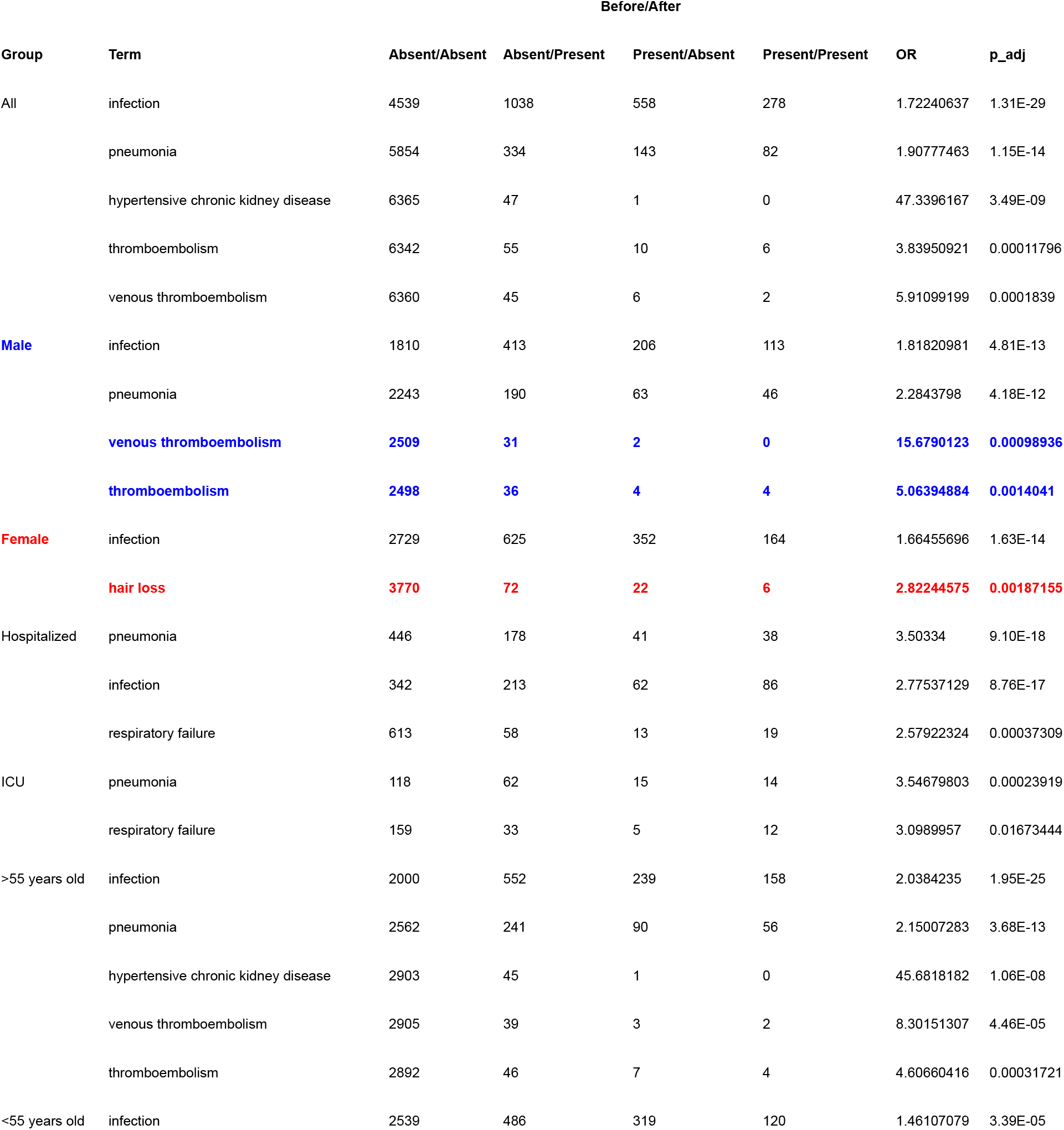
Signs and symptoms enriched after COVID-19 diagnosis compared to before.

| Group | Term | Before/After |  |  |  | OR | p_adj |
| --- | --- | --- | --- | --- | --- | --- | --- |
|  |  | Absent/Absent | Absent/Present | Present/Absent | Present/Present |  |  |
| All | infection | 4539 | 1038 | 558 | 278 | 1.72240637 | 1.31E-29 |
|  | pneumonia | 5854 | 334 | 143 | 82 | 1.90777463 | 1.15E-14 |
|  | hypertensive chronic kidney disease | 6365 | 47 | 1 | 0 | 47.3396167 | 3.49E-09 |
|  | thromboembolism | 6342 | 55 | 10 | 6 | 3.83950921 | 0.00011796 |
|  | venous thromboembolism | 6360 | 45 | 6 | 2 | 5.91099199 | 0.0001839 |
| Male | infection | 1810 | 413 | 206 | 113 | 1.81820981 | 4.81E-13 |
|  | pneumonia | 2243 | 190 | 63 | 46 | 2.2843798 | 4.18E-12 |
|  | venous thromboembolism | 2509 | 31 | 2 | 0 | 15.6790123 | 0.00098936 |
|  | thromboembolism | 2498 | 36 | 4 | 4 | 5.06394884 | 0.0014041 |
| Female | infection | 2729 | 625 | 352 | 164 | 1.66455696 | 1.63E-14 |
|  | hair loss | 3770 | 72 | 22 | 6 | 2.82244575 | 0.00187155 |
| Hospitalized | pneumonia | 446 | 178 | 41 | 38 | 3.50334 | 9.10E-18 |
|  | infection | 342 | 213 | 62 | 86 | 2.77537129 | 8.76E-17 |
|  | respiratory failure | 613 | 58 | 13 | 19 | 2.57922324 | 0.00037309 |
| ICU | pneumonia | 118 | 62 | 15 | 14 | 3.54679803 | 0.00023919 |
|  | respiratory failure | 159 | 33 | 5 | 12 | 3.0989957 | 0.01673444 |
| >55 years old | infection | 2000 | 552 | 239 | 158 | 2.0384235 | 1.95E-25 |
|  | pneumonia | 2562 | 241 | 90 | 56 | 2.15007283 | 3.68E-13 |
|  | hypertensive chronic kidney disease | 2903 | 45 | 1 | 0 | 45.6818182 | 1.06E-08 |
|  | venous thromboembolism | 2905 | 39 | 3 | 2 | 8.30151307 | 4.46E-05 |
|  | thromboembolism | 2892 | 46 | 7 | 4 | 4.60660416 | 0.00031721 |
| <55 years old | infection | 2539 | 486 | 319 | 120 | 1.46107079 | 3.39E-05 |

**Figure 2.**
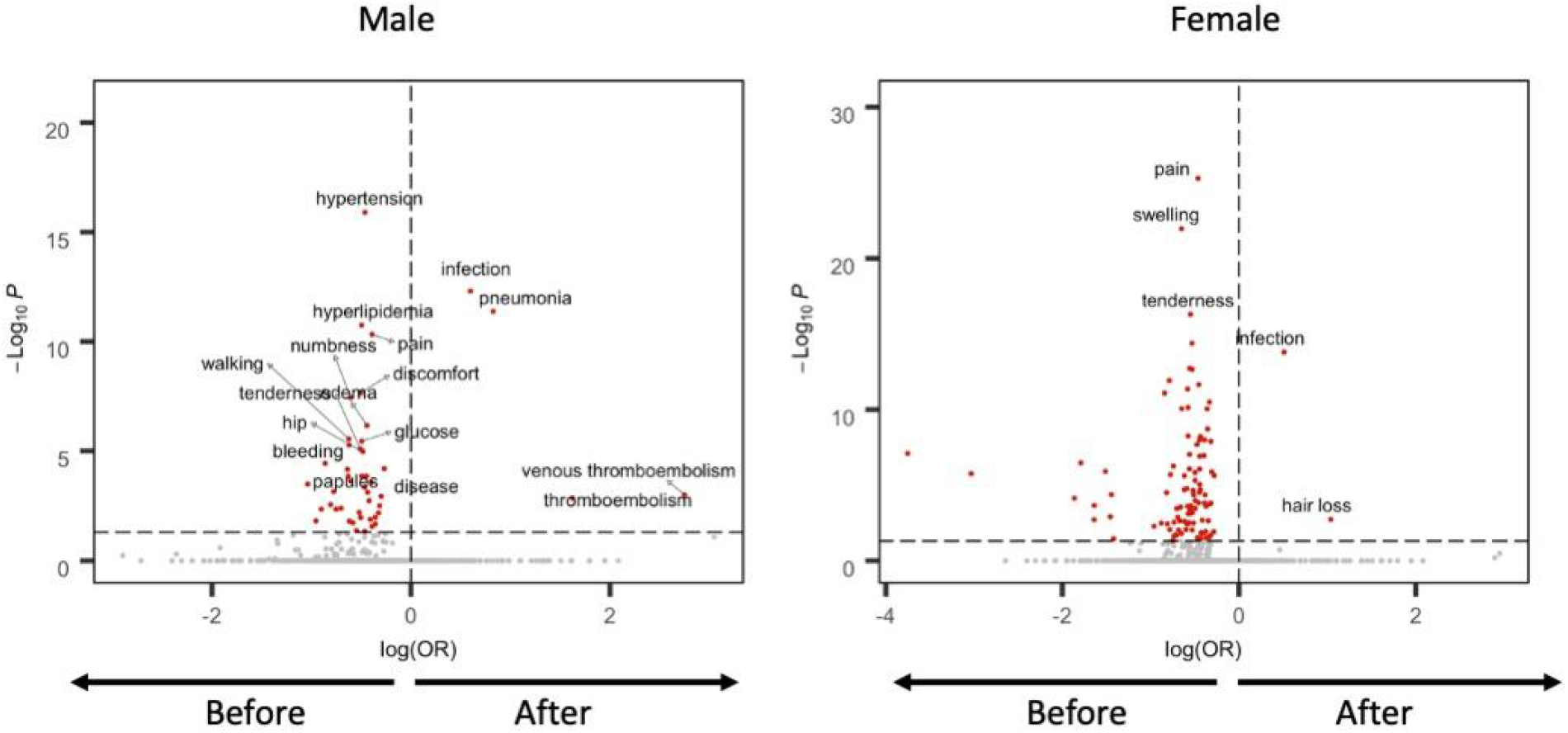
Males and females have distinct signs and symptoms in long COVID. Volcano plot displaying odds ratios (OR) of the change in prevalence before vs after disease for each sign and symptom identified from patient charts of male (left) and female (right) COVID-19 patients. P-values calculated by McNemar test and adjusted for multiple comparisons using Bonferroni correction. Significant (p<0.05) signs and symptoms indicated in red.

**Figure 3.**
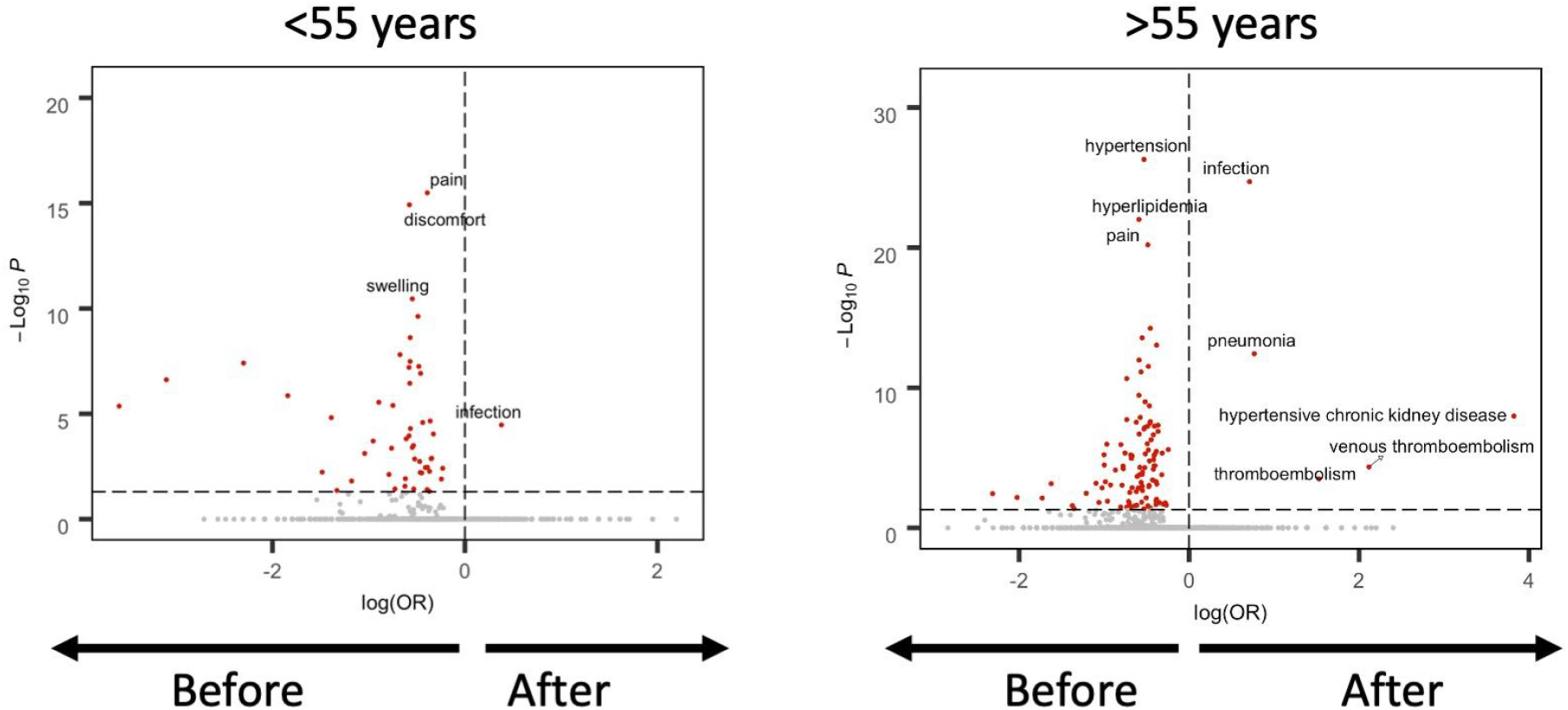
Patients of different ages have distinct signs and symptoms in long COVID. Volcano plots displaying odds ratios (OR) of the change in prevalence before vs after disease for each sign and symptom identified from patient charts of patients under 55 years old (left) and patients above 55 years old (right) with COVID-19. P-values calculated by McNemar test and adjusted for multiple comparisons using Bonferroni correction. Significant (p<0.05) signs and symptoms indicated in red.

**Figure 4.**
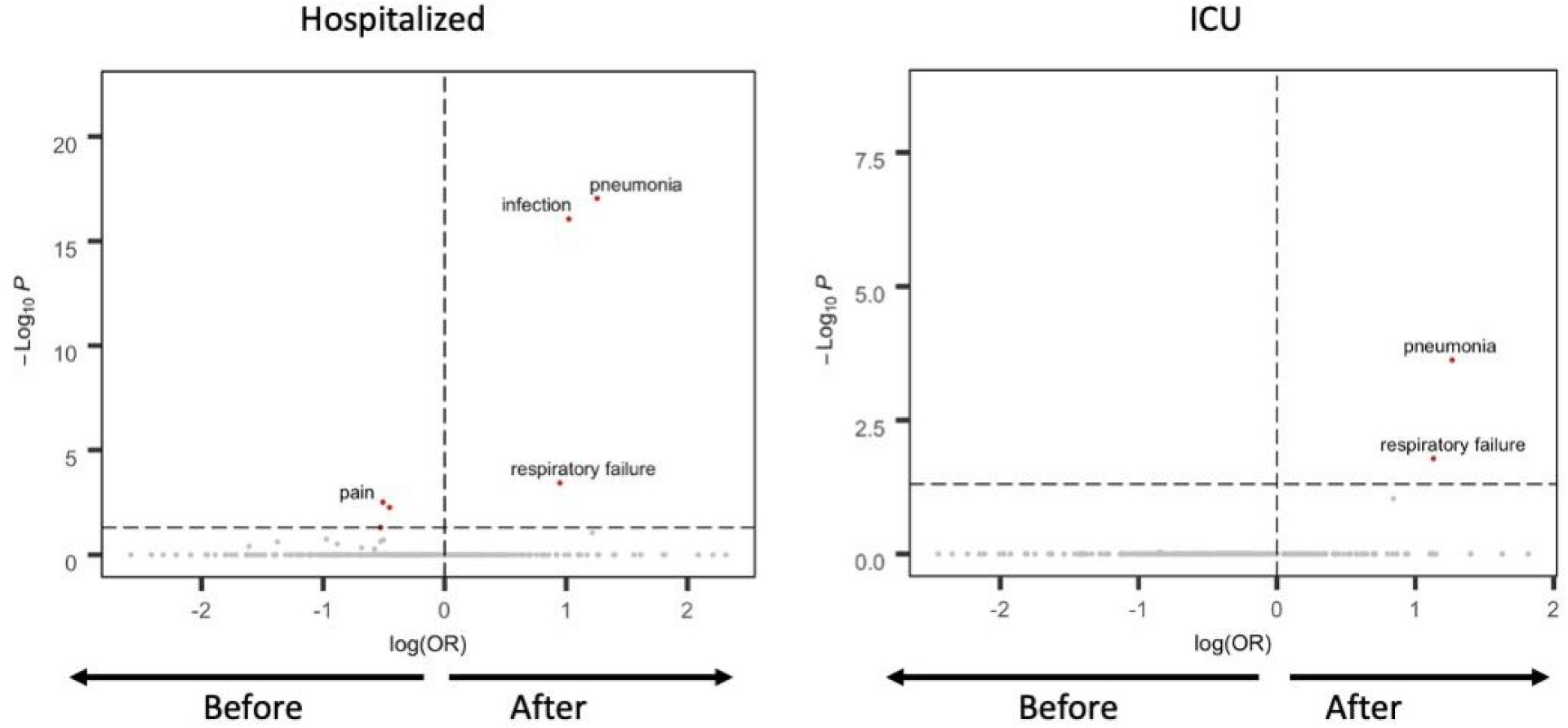
Hospitalized and patients admitted to the ICU have similar signs and symptoms in long COVID. Volcano plot displaying odds ratios (OR) of the change in prevalence before vs after disease for each sign and symptom identified from patient charts of hospitalized (left) and ICU-admitted (right) COVID-19 patients. P-values calculated by McNemar test and adjusted for multiple comparisons using Bonferroni correction. Significant (p<0.05) signs and symptoms indicated in red.

Although we focused on notes in the 3-6 months after COVID-19 diagnosis, it is important to realize that the documentation of a phenotype in this time interval does not necessarily imply that it actually occurred during this interval. For example, the enrichment of infection and pneumonia in the post-COVID period likely reflects a discussion of the prior COVID-19 infection rather than the development of a new infection. To distinguish such signals that may reflect phenotypes which occurred in the peri-COVID window from those that truly emerge in the post-COVID period, we examined the cumulative incidence of each phenotype found to be enriched in the post-COVID period as a function of time relative to COVID diagnosis (Figure 5). Indeed, we observe a distinct acceleration in occurrences of “hair loss” among females and “thromboembolism”/”venous thromboembolism” among males in the post-COVID period, whereas this was not observed for “pneumonia”, “hypertensive chronic kidney disease”, and “respiratory failure”. Taken together, these data suggest that hair loss and thromboembolism warrant further study as potentially unique features of long COVID in females and males, respectively.

**Figure 5.**
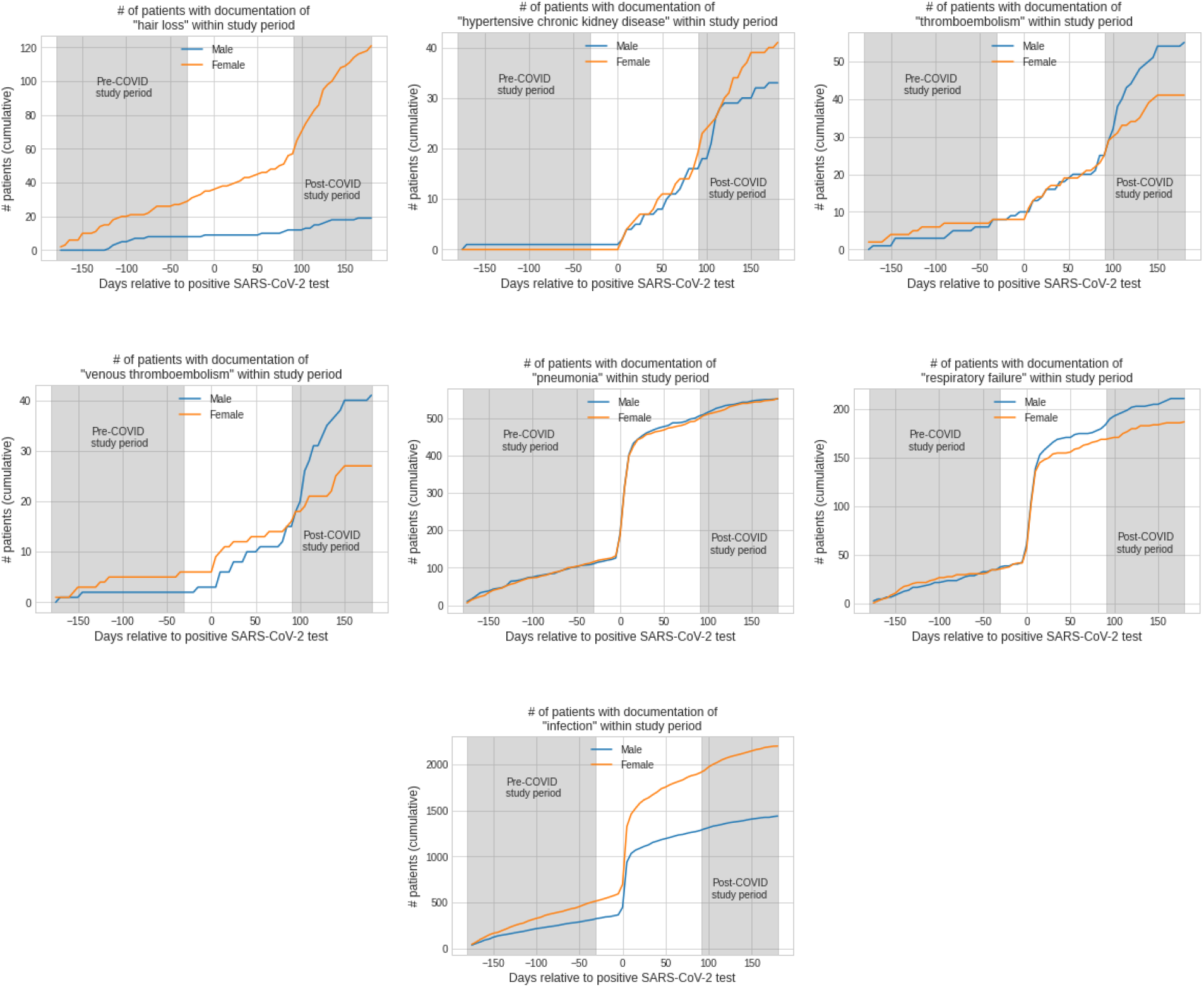
Cumulative patient counts of possible long-COVID symptoms and diseases relative to COVID-19 diagnosis during the study period. Patients are counted as symptoms and diseases are attributed to them in physician notes during the study period (−180 days to +180 days relative to COVID-19 diagnosis).

## Discussion

Through augmented curation analysis of over sixty thousand COVID-19 patient charts, we have identified a range of phenotypes, including type 2 diabetes, thromboembolism, and hair loss, as possible long-term associations with COVID-19 diagnosis. By comparing COVID-19 patients before and after diagnosis, we have unbiasedly characterized signs and symptoms both due to disease onset, and signs and symptoms specific to a given respiratory infection.

Early investigations of long COVID have provided helpful preliminary characterization of the sequelae of signs and symptoms that arise months after COVID-19 diagnosis. Among signs and symptoms reported in the months after COVID-19 diagnosis, the most common associations with long COVID include fatigue, cough, myalgia, headache, memory problems, and anosmia. Notably, these analyses have relied on structured patient data or self-reported symptoms, and are therefore limited to a finite set of biased variables. The present analysis of physician-documented diseases, signs, and symptoms in unstructured EHR data provides an unbiased approach to identify signs and symptoms associated with long COVID. Reassuringly, our findings corroborate early studies of the signs and symptoms of long COVID, including headache and hair loss, emphasizing the importance of monitoring for these in the months after COVID-19 diagnosis. We also report several new associations with long COVID, including hypertensive chronic kidney disease and thromboembolism, which may warrant further investigation and validation using structured patient data. Overall, these differences between our analysis using augmented curation and previous analyses of long COVID highlight the need for a deeper understanding of the differences between information captured in unstructured compared to structured health data.

The approach to automatically curating physician notes taken here has limitations. First, we curate whether or not a physician is ascribing a diagnosis or symptom to a patient independent of the acuity of the diagnosis. Documentation of a prior or historical illness is not differentiated from a newly documented illness. The within-patient comparison of post-COVID symptoms and diseases to pre-COVID symptoms and diseases mitigates this effect, but it is important to note that the pre-COVID period covers only the 1-6 months prior to COVID-19 diagnosis. Furthermore, though the inclusion criteria for the study require at least one symptom or disease documented in both the pre-COVID and post-COVID period, the type of clinical document(s) that this symptom or disease may be identified in is left unrestricted. Thus, comprehensive patient medical histories may not be captured in the pre-COVID period.

As the COVID-19 pandemic continues, there is mounting concern over its long-term consequences. Although our understanding of the acute manifestations of COVID-19 has improved, disease presentation and long-term effects remain relatively uncharacterized due to the paucity of long-term patient data. Augmented curation allows us to begin to address this shortcoming by unbiasedly extracting key word associations from tens of thousands of patient charts. Here we have identified a range of phenotypes, including thromboembolism and hypertensive chronic kidney disease, as possible long-term associations with COVID-19 diagnosis. These findings compel targeted investigations into what may be persistent coagulopathic phenotypes following SARS-CoV-2 infection.

## Methods

### Study design

This retrospective study was comprised of patients who presented to the Mayo Clinic Health System (including tertiary medical centers in Minnesota, Arizona, and Florida) and received at least one positive SARS-CoV-2 PCR test since the start of the COVID-19 pandemic and further limited to those that have had at least one symptom or disease attributed to them in unstructured clinical documentation during the 1 to 5 months prior to COVID positive test and 3 to 6 months post COVID positive test (n = 6,432). COVID_pos_ patients were identified via a positive PCR test result.

### Natural Language Processing / augmented curation of electronic health records (EHRs)

We used previously developed and detailed state-of-the-art BERT-based neural networks^13^ to rapidly curate clinical notes that were authored within 6 months of and COVID-19 diagnoses. Specifically, the model extracts sentences containing clinical phenotypes and symptoms and classifies their sentiment into the following categories: Yes (confirmed clinical manifestation or diagnosis), No (ruled out clinical manifestation or diagnosis), Maybe (possibility of clinical manifestation or diagnosis), and Other (alternate context, e.g., family history of disease). The neural networks are pre-trained on 3.17 billion tokens from the biomedical and computer science domains (SciBERT)^14^ and subsequently trained using 18,490 sentences and approximately 250 phenotypes with an emphasis on cardiovascular, pulmonary, and metabolic phenotypes. It achieves 93.6% overall accuracy and over 95% precision and recall for both “Yes” and “No” sentiment classification.

### Statistical analyses

To compute the statistical significance of enrichment of a specific condition following a COVID-19 diagnosis, McNemar’s test was employed. This test is specifically designed for matched pairs with binary outcomes (here, the absence or presence of symptom or disease in time period of interest) and evaluates the null hypothesis that the probability of a specific condition occurring following a COVID-19 diagnosis is equal to the probability of the aforementioned condition occurring before said diagnosis. In this analysis we used the exact version of the McNemar test to compute the p-value for the null hypothesis. We computed the Risk Difference (the difference between the probability of a condition occurring following the diagnosis and the probability of the same condition occurring before the diagnosis), the Risk Ratio (the ratio of the probability of a condition occurring following the diagnosis to the probability of the same condition occurring before the diagnosis), and the Odds Ratio (the ratio of the odds of a condition occurring following the diagnosis to the odds of the same condition occurring before the diagnosis), as well as their respective 95% Wald Confidence Intervals. All multiple hypothesis tests were Bonferroni corrected.

### Institutional Review Board (IRB)

This is a retrospective study of individuals who underwent polymerase chain reaction (PCR) testing for suspected SARS-CoV-2 infection at the Mayo Clinic and hospitals affiliated with the Mayo Clinic Health System. This study was reviewed by the Mayo Clinic Institutional Review Board (IRB) and determined to be exempt from the requirement for IRB approval (45 CFR 46.104d, category 4). Subjects were excluded if they did not have a research authorization on file.

### Patient and public involvement

The development of the research question and outcome measures was informed by prior literature and information from the Centers for Disease Control and Prevention (CDC) on risk factors for severe COVID-19 illness. No patients were involved in the design of the study, but physicians from the Mayo Clinic who are involved with the COVID-19 research taskforce and the clinical care for COVID-19 patients were involved with the study design and execution.

## Data Availability

After publication, the data will be made available to others upon reasonable requests to the corresponding author. A proposal with detailed description of study objectives and statistical analysis plan will be needed for evaluation of the reasonability of requests. Deidentified data will be provided after approval from the corresponding author and the Mayo Clinic standard IRB process for such requests.

## Conflict of Interest Statement

ADB is a consultant for Abbvie, is on scientific advisory boards for nference and Zentalis, and is founder and President of Splissen therapeutics. One or more of the investigators associated with this project and Mayo Clinic have a Financial Conflict of Interest in technology used in the research and that the investigator(s) and Mayo Clinic may stand to gain financially from the successful outcome of the research. This research has been reviewed by the Mayo Clinic Conflict of Interest Review Board and is being conducted in compliance with Mayo Clinic Conflict of Interest policies. The authors from nference have financial interests in nference.

## Notes

### Competing Interest Statement

The authors have declared no competing interest.

### Funding Statement

No external funding supported this work.

### Author Declarations

This study was reviewed and approved by the Mayo Clinic Institutional Review Board as a minimal risk study (IRB 20-003278, 'Study of COVID-19 patient characteristics with augmented curation of Electronic Health Records (EHR) to inform strategic and operational decisions'). Subjects were excluded if they did not have a research authorization on file.

### Summary of Updates

Substantially updated the manuscript by curating an additional 6 months worth of data, resulting in a 20-fold expansion of the COVID cohort size. Focused the paper on the within COVID analysis rather than COVID vs. influenza.

## References

1. Yelin, D. et al. Long-term consequences of COVID-19: research needs. Lancet Infect. Dis. 20, 1115–1117 (2020).

2. Mahase, E. Covid-19: What do we know about “long covid”? BMJ 370, m2815 (2020).

3. Tenforde, M. W. et al. Symptom Duration and Risk Factors for Delayed Return to Usual Health Among Outpatients with COVID-19 in a Multistate Health Care Systems Network - United States, March-June 2020. MMWR Morb. Mortal. Wkly. Rep. 69, 993–998 (2020).

4. Carfì, A., Bernabei, R., Landi, F. & Gemelli Against COVID-19 Post-Acute Care Study Group. Persistent Symptoms in Patients After Acute COVID-19. JAMA 324, 603–605 (2020).

5. Zhao, Y.-M. et al. Follow-up study of the pulmonary function and related physiological characteristics of COVID-19 survivors three months after recovery. EClinicalMedicine 25, 100463 (2020).

6. Abbott, A. Thousands of people will help scientists to track the long-term health effects of the coronavirus crisis. Nature 582, 326 (2020).

7. Wagner, T. et al. Augmented curation of clinical notes from a massive EHR system reveals symptoms of impending COVID-19 diagnosis. eLife 9, (2020).

8. Shi, L. et al. Prevalence of and Risk Factors Associated With Mental Health Symptoms Among the General Population in China During the Coronavirus Disease 2019 Pandemic. JAMA Netw. Open 3, e2014053 (2020).

9. Taquet, M., Luciano, S., Geddes, J. R. & Harrison, P. J. Bidirectional associations between COVID-19 and psychiatric disorder: retrospective cohort studies of 62?354 COVID-19 cases in the USA. Lancet Psychiatry (2020) doi:10.1016/S2215-0366(20)30462-4.

10. Pawlowski, C. et al. Inference from longitudinal laboratory tests characterizes temporal evolution of COVID-19-associated coagulopathy (CAC). eLife 9, (2020).

11. Puntmann, V. O. et al. Outcomes of Cardiovascular Magnetic Resonance Imaging in Patients Recently Recovered From Coronavirus Disease 2019 (COVID-19). JAMA Cardiol. 5, 1265–1273 (2020).

12. Gklinos, P. Neurological manifestations of COVID-19: a review of what we know so far. J. Neurol. 267, 2485–2489 (2020).

13. Devlin, J., Chang, M.-W., Lee, K. & Toutanova, K. BERT: Pre-training of Deep Bidirectional Transformers for Language Understanding. in Proceedings of the 2019 Conference of the North American Chapter of the Association for Computational Linguistics: Human Language Technologies, Volume 1 (Long and Short Papers) 4171–4186 (Association for Computational Linguistics, 2019). doi:10.18653/v1/N19-1423.

14. Beltagy, I., Lo, K. & Cohan, A. SciBERT: A Pretrained Language Model for Scientific Text. in Proceedings of the 2019 Conference on Empirical Methods in Natural Language Processing and the 9th International Joint Conference on Natural Language Processing (EMNLP-IJCNLP) 3615–3620 (Association for Computational Linguistics, 2019). doi:10.18653/v1/D19-1371

